# A method for measuring mitochondrial DNA copy number in pediatric populations

**DOI:** 10.1101/2024.03.19.24304372

**Authors:** Simran Maggo, Liam Y. North, Aime Ozuna, Dejerianne Ostrow, Grajeda I Yander, Hakimjavadi Hesmedin, Jennifer A. Cotter, Alexander R. Judkins, Pat Levitt, Xiaowu Gai

## Abstract

The mitochondrion is a multifunctional organelle that modulates multiple systems critical for homeostasis during pathophysiological stress. Variation in mitochondrial DNA (mtDNA) copy number (mtDNAcn), a key mitochondrial change associated with chronic stress, is an emerging biomarker for disease pathology and progression. mtDNAcn can be quantified from whole blood samples using qPCR to determine the ratio of nuclear DNA to mtDNA. However, the collection of blood samples in pediatric populations, particularly in infants and young children, can be technically challenging, yield much smaller volume samples, and can be distressing for the patients and their caregivers. Therefore, we have validated a mtDNAcn assay utilizing DNA from simple buccal swabs (Isohelix SK-2S) and report here it’s performance in specimens from infants (age = <12 months). Utilizing qPCR to amplify ∼200bp regions from two mitochondrial (*ND1, ND6*) and two nuclear (*BECN1, NEB*) genes, we demonstrated absolute (100%) concordance with results from low-pass whole genome sequencing (lpWGS). We believe that this method overcomes key obstacles to measuring mtDNAcn in pediatric populations and creates the possibility for development of clinical assays to measure mitochondrial change during pathophysiological stress.

## 1 Introduction

The mitochondrion is an essential organelle producing cellular energy through oxidative metabolism. In addition to being the “powerhouse” of the cell, mitochondria are increasingly recognized for their roles in a broad range of cellular processes. Damage-associated molecular patterns (DAMPs) are pathophysiological changes that occur on the cellular level that can be mediated by the release of mitochondrial DNA and mitochondrial double stranded RNA into the cytoplasm during mitophagy (1, 2). This may lead to increased oxidative stress and changes in nuclear gene expression, triggering inflammation-related signaling pathways and systemic inflammatory responses (3–5).

One of the most well-studied associations between mtDNAcn and pathology is in cancer (6). Increased mtDNAcn has been observed in various cancer types, including breast, lung, and colorectal cancer. Known as mitochondrial biogenesis, this is thought to support the high energy demands of rapidly dividing cancer cells, but may also contribute to chemoresistance, making it a potential therapeutic target (7–9). Diseases may also be characterized by a reduced mtDNAcn. Mitochondrial diseases often involve mutations in nuclear or mitochondrial genes that disrupt mtDNA maintenance affecting energy production and causing a wide range of symptoms, such as muscle weakness, neurological deficits, and organ dysfunction (1, 10, 11). Other diseases associated with changes in mtDNAcn include neurodegenerative and cardiovascular diseases, and metabolic disorders including type 2 diabetes and obesity (12–14).

Chronic stress, a persistent state of emotional and psychological strain, has been closely linked with changes in mtDNAcn and altered mitochondrial functioning in pre-clinical models of early adversity and in human adults (15–17). Disordered mitochondrial function may be an important contributor to the molecular mechanisms of human disease, including several mental and physical health conditions. In chronic stress, dysregulation of the hypothalamic-pituitary-adrenal (HPA) axis and increased cortisol can affect mitochondrial function and mtDNAcn (18, 19). These changes highlight the need for a reliable means of assessing mtDNAcn.

A variety of methods have been described to quantify mtDNAcn, beginning with use of a combination of restriction enzymes and southern blotting to evaluate the intensity of nuclear to mitochondrial DNA probes (20). Molecular methods including PCR, qPCR, digital PCR and genomic methods including whole exome sequencing (WES) and whole genome sequencing (WGS) have all been used to establish a relative mtDNAcn value by comparing nuclear to mitochondrial intensity, CT values, copies, number of reads, respectively (3, 21, 22). PCR based approaches are broadly cost effective for implementation of routine clinical testing. While WES/WGS are becoming more cost-effective and are used in some specialized settings, they remain impractical for routine clinical testing of mtDNAcn. However, given the small size of the mitochondrial genome, low pass whole genome sequencing (lpWGS) at a read depth of 0.5 – 3x provides ∼50X mitochondrial genome coverage. This is sufficient for quantifying mtDNAcn and is significantly less expensive than standard WES/WGS in settings with the capacity for clinical genomic testing (23).

To obtain sufficient volume and quality of DNA for mtDNAcn testing, conventional methods utilize venipuncture, from which up to 10mLs of whole blood is collected (24). However, this exceeds the recommended volume limits for infants and children (25), and venipuncture can be technically challenging and can generate anxiety and discomfort in pediatric populations (26–28). Therefore, we utilized buccal swabs (Isohelix SK-2S buccal swab kit and BuccalFix buffer BFX25 to stabilize DNA and RNA) for specimen collection as it was minimally invasive, simple to administer and cost effective. Using DNA extracted from these swabs, we developed a mtDNAcn qPCR assay and evaluated its performance in comparison to a commercially available mtDNAcn qPCR kit (NovaQUANT Human Mitochondrial to Nuclear DNA ratio kit, Millipore Sigma, USA) and lpWGS.

## 2 Materials and Methods

### 2.1 Participants and study design

Buccal swab samples collected through Children’s Hospital Los Angeles as part of the California Initiative to Advance Precision Medicine (CIAPM) - Adverse Childhood Experiences (ACEs) program (Scalable Measurement and Clinical Deployment of Mitochondrial Biomarkers of Toxic Stress – IRB# CHLA-21-00174) and the Your Baby: Healthy Development and Resiliency (IRB# CHLA-18-00547) studies were used for assay development. Participants’ or their legal representatives provided written consent to take part in the aforementioned studies. The ACEs study samples used in this study were from 164 mothers and their infants recruited to participate at the 6-month timepoint of this ongoing study that also includes a 12-month infant visit. The Your Baby study samples used in this study were from 19 mothers and 21 infants. A total of six samples collected as part of the CIAPM study were used to validate the commercial (samples a1-a3) and our own (samples b1-b3) qPCR based mtDNAcn assay. The commercial assay and lpWGS were performed on 40 samples (Your Baby study, 19 mothers and their 21 infants) to assess the concordance of these two techniques. However, further use of the commercial qPCR mtDNAcn assay was precluded by supply chain issues which limited the availability of kits (at the time of manuscript submission, the NovaQUANT assay is currently described as “limited availability” from Millipore Sigma). Therefore, we established the concordance of our assay (developed with CIAPM samples b1-b3) using three samples from the Your Baby study that were previously used for the commercial assay and lpWGS. We then performed our mtDNAcn qPCR assay on 164 mother and 164 infant ACEs study samples and, finally assessed concordance using lpWGS on a subset of 36 samples (18 mothers and their 18 infants) from the ACEs study. The workflows and study samples are summarized in **Figure 1**.

**Figure 1.**
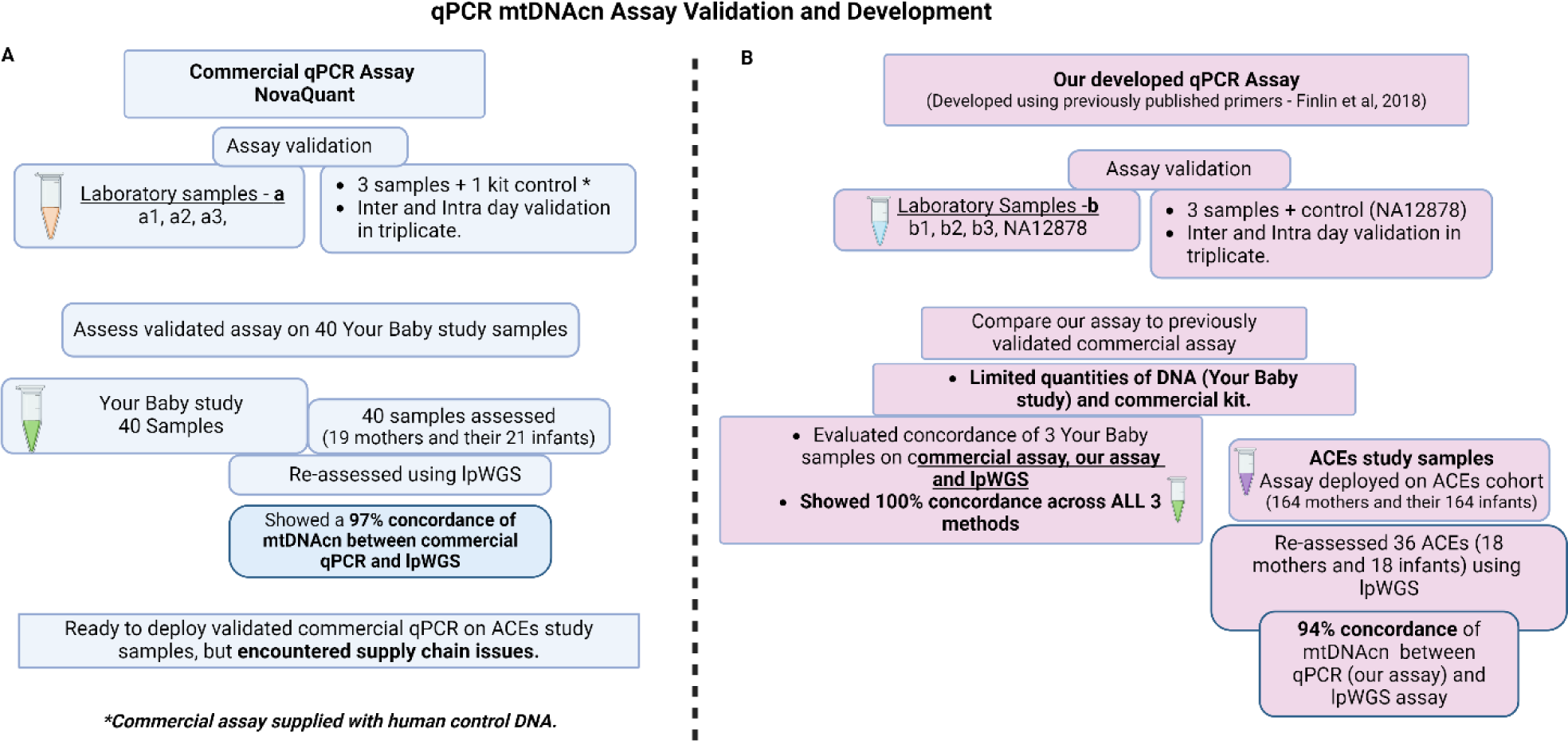
Study Design for qPCR mtDNA assay validation and development. (A) Commercial, NovaQUANT (Millipore Sigma, USA) mtDNAcn qPCR assay validation and study samples utilized; (B) our mtDNAcn qPCR assay development, validation and study samples utilized.

### 2.2 Specimen collection, DNA isolation and extraction

The IsoHelix SK-2S kit in combination with the BFX-25 stabilizing buffer can be used for the isolation of both DNA and RNA from a single buccal swab. Caregivers were instructed not to give infants food or milk for 60 minutes prior to specimen collection. Mothers utilized a gentle ‘bear hug technique’ to hold the infant while a member of the research staff swabbed the buccal mucosa of each cheek for 30 seconds (total of 60 seconds, timed), and then placed the swab into the collection tube. Maternal buccal swabs were self-collected during the same visit following an identical procedure. While some infants tolerated a shorter period of time (30-45 seconds), we found that moderate pressure applied during swabbing for any time between 30-60 seconds is the most important element for sufficient sample collection. No mother-infant pairs were excluded from the study due to inadequate swab times. Collection tubes received a 500 μL aliquot of BuccalFix buffer (BFX-25) and were inverted 10 times before being barcoded and stored at −80°C in the CHLA Pediatric Research Biorepository. DNA and RNA were extracted using the Maxwell RSC Blood DNA kit &RSC miRNA from Tissue and Plasma Kit (Promega) and stored at 4°C and −80°C, respectively, until required. DNA/RNA quality (nanodrop) and quantity (Qubit) was assessed prior to use.

### 2.3 mtDNAcn assays and lpWGS

NovaQUANT (Millpore Sigma, USA) is a qPCR assay that assesses relative mtDNAcn by comparing the ratio of mitochondrial to nuclear DNA using the Ct (ΔCt) levels of two mitochondrial genes (ND1 and ND6) and two nuclear genes (BECN1 and NEB). Real-time PCR of 4 targets (2 mitochondrial, 2 nuclear) was performed following the manufacturer’s protocol. All samples were run in triplicate (DNA – 2ng/replicate), including nuclease-free water as a no template control (NTC). For analysis, we used the manufacturer’s recommended method in which relative mtDNAcn was calculated by assessing the ratio of mtDNA/nDNA for the MT-ND1 and MT-ND4, by the geometric mean of BECN1 and NEB1.

Due to supply chain issues that limited availability of the commercial assay, we designed a mtDNAcn qPCR assay utilizing prior published primers and mitochondrial gene target normalization against nuclear genes (29) (Table1). The Ct (ΔCt) levels of two mitochondrial genes (ND1 and ND6) and two nuclear genes (BECN1 and NEB) were assessed for each DNA sample. A reference DNA-sample (NA12878) which has been broadly used in validating many genomic assays was included in all runs. Real-time PCR of the four target genes was performed on DNA obtained from the buccal swab specimens using the following cycling parameters: 95°C for 10 minutes; 40 cycles of 95°C for 15 seconds, 60°C for 60 seconds on the StepOnePlus™ Real-Time PCR System (Applied Biosystems, USA) and using RT² SYBR Green ROX FAST Master mix (Qiagen, USA) consisting of HotStart DNA taq polymerase, nucleotides, ROX as the reference dye, and SYBR Green dye. All samples were run in triplicate (DNA – 2ng/replicate), including nuclease-free water as a no template control (NTC). Once normalized, the mitochondrial gene target Ct value was analyzed using the 2-Delta-Delta method (30) to convert the Ct value to a linear form for analysis of gene expression and calculate the relative change in mitochondrial copy number. Briefly, Cts from the ND1 gene are subtracted from that of the BECN1 gene to obtain ΔCt1 (ΔCt1 = Ct^Nuc1^-Ct^Mito1^), while ND6 Ct is subtracted from NEB Ct to obtain ΔCt2 (ΔCt2 = Ct^Nuc2^-Ct^Mito2^), then copy numbers are calculated based on the ΔCt of the matched mitochondrial to nuclear DNA Cts (N = 2^ΔCt^). The average of the two copy number results provides the final relative mtDNAcn per sample.

**Table 1.**
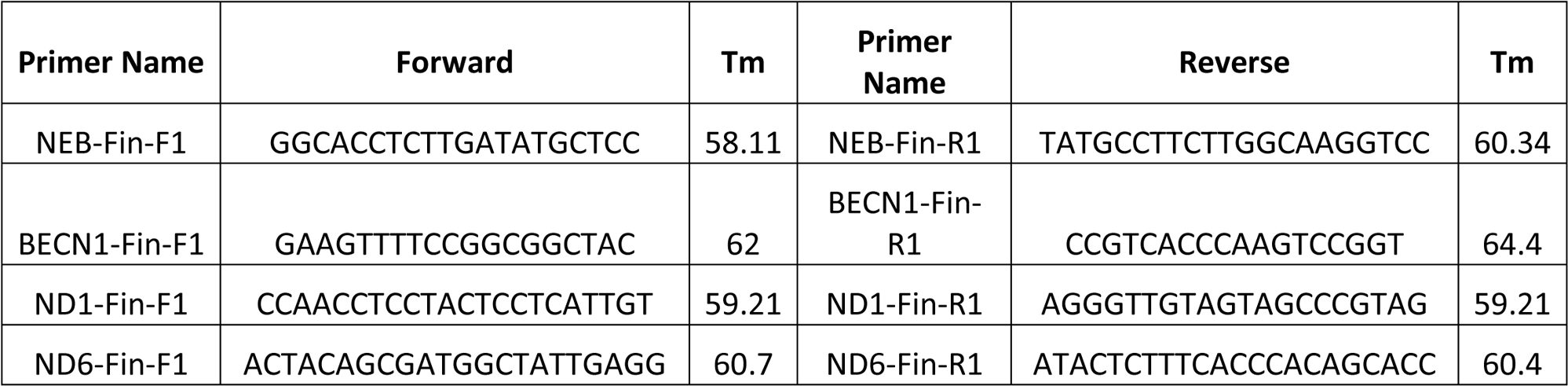
Primers utilized.

We performed lpWGS to evaluate the performance of both the commercial and our own mtDNAcn assay (**Figure 1**). DNA (5ng) obtained from a subset of the same buccal swab specimens was used to prepare NGS libraries using xGen™ cfDNA & FFPE DNA Library Prep (Integrated DNA Technologies Coralville, IA). NGS libraries were barcoded, pooled and sequenced on Illumina NextSeq 500 at 2×100bp to an average read-depth ranging from 0.5 – 1X. Relative mtDNAcn was then calculated using the formula mtDNAcn= (Average mitochondrial genome coverage/Average autosomal chromosome coverage)*2, as previously described (31).

We assessed the performance of our mtDNAcn assay by three-way comparison to the commercial assay and lpWGS results obtained from three samples using DNA (2ng/replicate) obtained from a subset of the same buccal swab specimens from the Your Baby study. We were only able to perform this 3-way comparison on this subset of samples due to (1) limited amounts of DNA remaining and (2) limited quantities of the commercial assay.

### 2.4 Data analysis

Correlation analysis and graphing were done using GraphPad Prism (version 9.5.1) and R (version 4.3.1) running in RStudio. Data are presented as means with error bars depicting standard deviation (SD) unless otherwise specified.

## 3 Results

### 3.1 Commercial assay validation and assessment

We initially assessed buccal swab specimens from the Your Baby study comprised of 19 mothers and 21 infants. As shown in **Figure 2**, the IsoHelix SK-2S kit in combination with the BFX-25 stabilizing buffer produced high-quality DNA and RNA as assessed by 260/280 and 260/230 ratios. The collection method yielded a range of 20-2000ng DNA across infant samples, and 200-4000ng DNA across maternal samples (**Figure 2C**). We were able to elute an average of 600ng of DNA and RNA in a 25uL volume, providing sufficient nucleic acid concentrations for multiple assays on each sample (new **Figures 2C and 2D**). Prior to assessing mtDNAcn in the Your Baby study samples, we conducted an inter and intra-day assay validation of the commercial NovaQUANT mtDNA qPCR assay utilizing DNA obtained from three specimens (a1, a2 and a3). Inter-day variability was determined by running a1, a2 and a3 in triplicate on consecutive days; intra-day variability was determined by running the same samples in triplicate a second time on the same plate. (**Supplementary Figures s1 – s3**).

**Figure 2.**
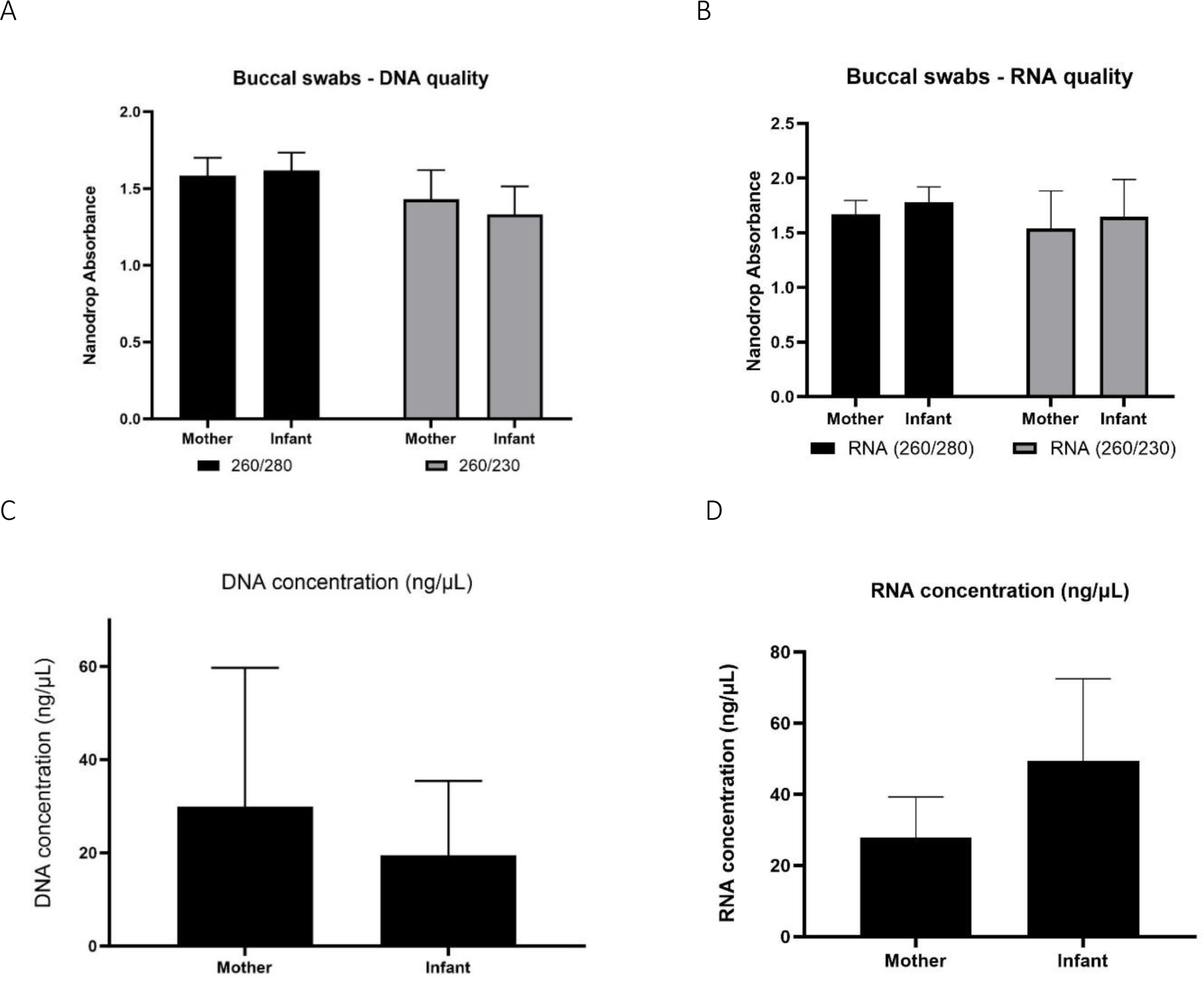
Nucleic acid collection results. Assessment of DNA and RNA quality (panels A and B) and quantity (panels C and D) of the IsoHelix SK-2S kit in combination with the BFX-25 stabilizing buffer on 19 mother and 21 infant samples from the Your Baby study.

We then used the commercial assay to determine mtDNAcn for Your Baby study samples utilizing DNA available for 19 mothers and 21 infants. Our results shown in **Figure 3A** illustrate the mean mtDNAcn level was higher in infants (229.2 ± 66.4 S.D) than maternal samples (136.7 ± 41.8 S.D) (**Figure 3A**). The Your Baby samples showed a 97% concordance (**Figure 3B**) between the commercial mtDNAcn qPCR assay and lpWGS results.

**Figure 3.**
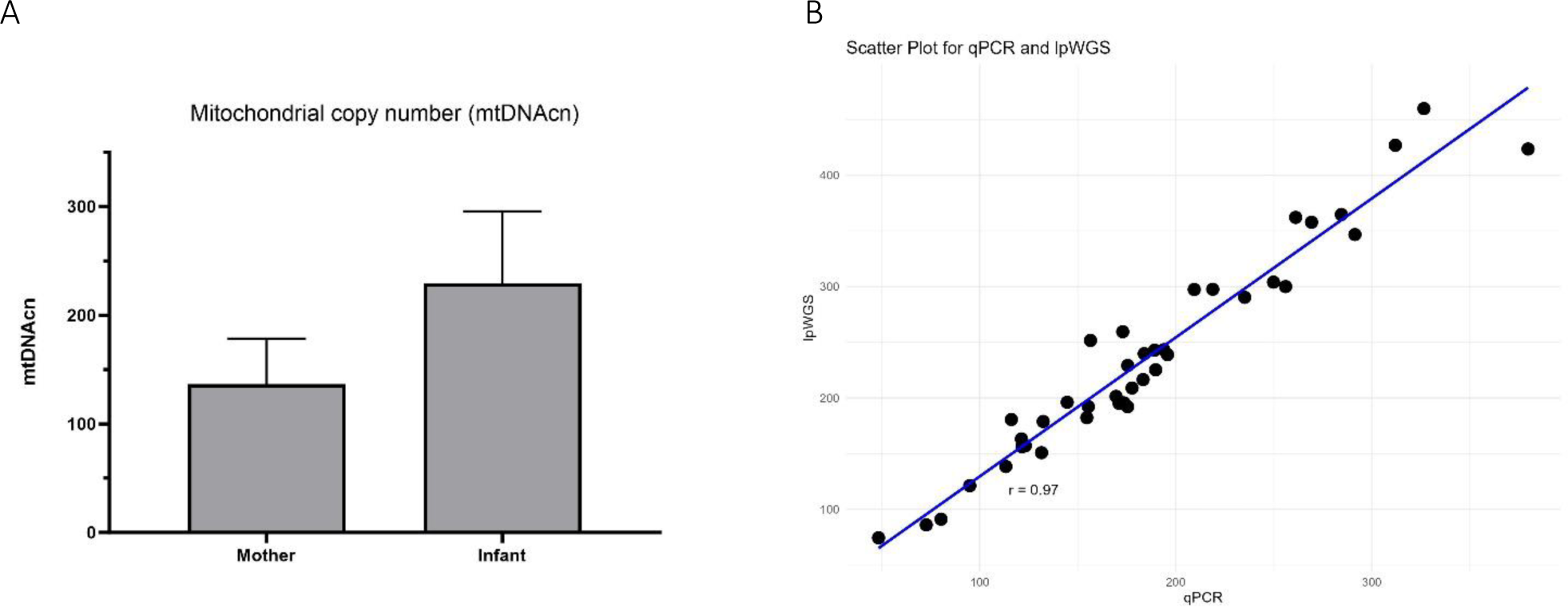
mtDNAcn results for commercial assay and concordance with lpWGS. Mother and infant mtDNAcn assessment using the commercial assay (panel A). Strong (Pearson r= 0.97) concordance of the commercial qPCR assay and lpWGS for mtDNAcn assessment (panel B).

### 3.2 Validation and performance characterization of our qPCR assay

We developed our own custom mtDNAcn qPCR assay combining methods from the commercial assay and published primers as previously described. We performed inter- and intra-day validation for our assay as described previously (**Supplementary Figures s4-s6**). Using three samples from the Your Baby Study, we performed a three-way comparison of our assay, the commercial assay and lpWGS assay, and showed a 100% concordance (Pearson r of 1) for all three assays (**Figure 4**).

**Figure 4.**
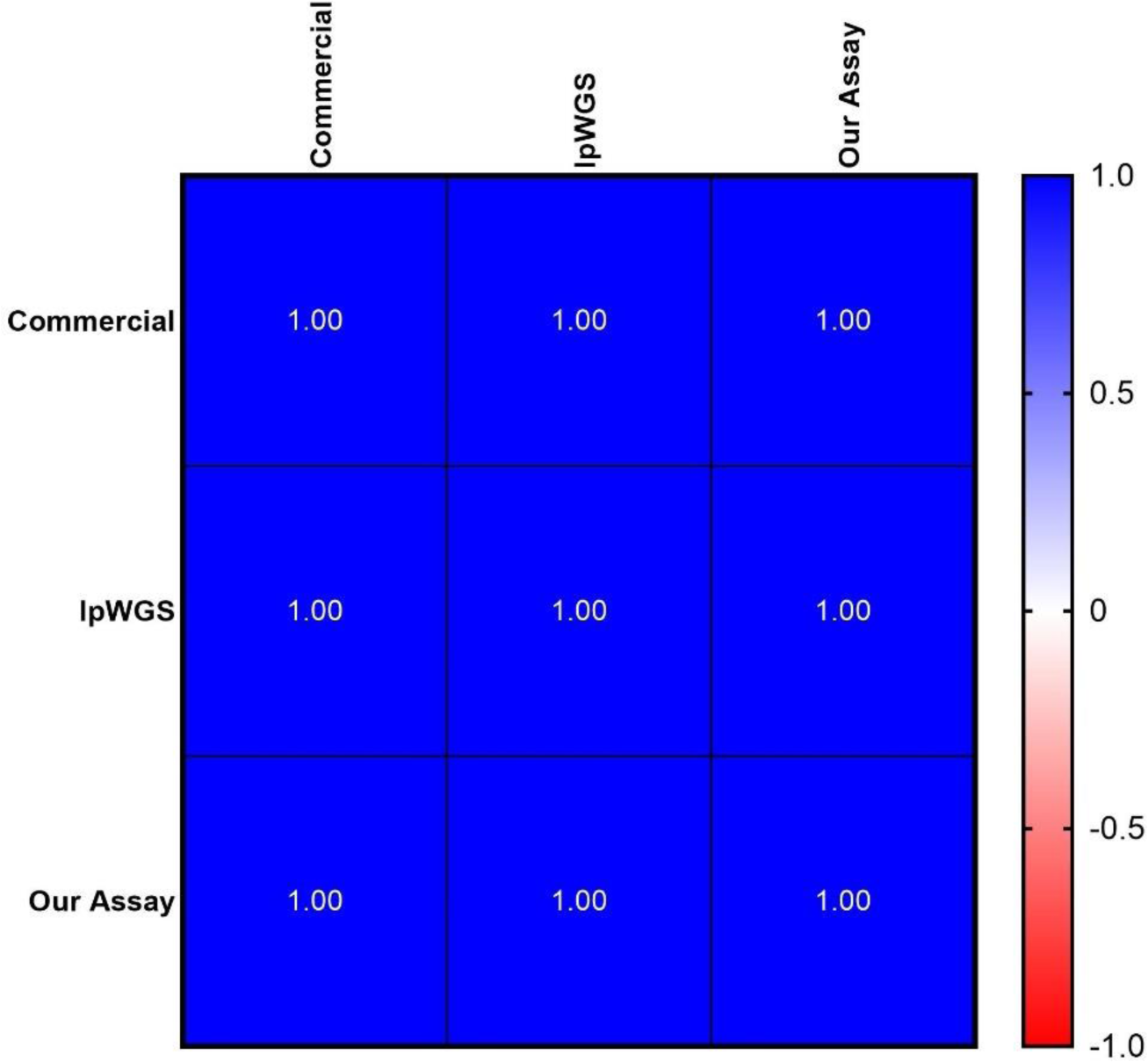
Concordance of mtDNAcn between commercial, lpWGS and our own assays. High (100%) concordance, **depicted as Pearson’s r values**, of all three methodologies used to assess mtDNAcn.

Our own assay then was utilized to evaluate DNA from 164 mothers and 164 six-month old infant samples collected for the ACEs study. **Figure 5** shows DNA quality, quantity and mtDNAcn from this cohort. In addition, we showed a high concordance between our assay and lpWGS on a subset of 36 ACEs samples (Pearson r = 0.94).

**Figure 5.**
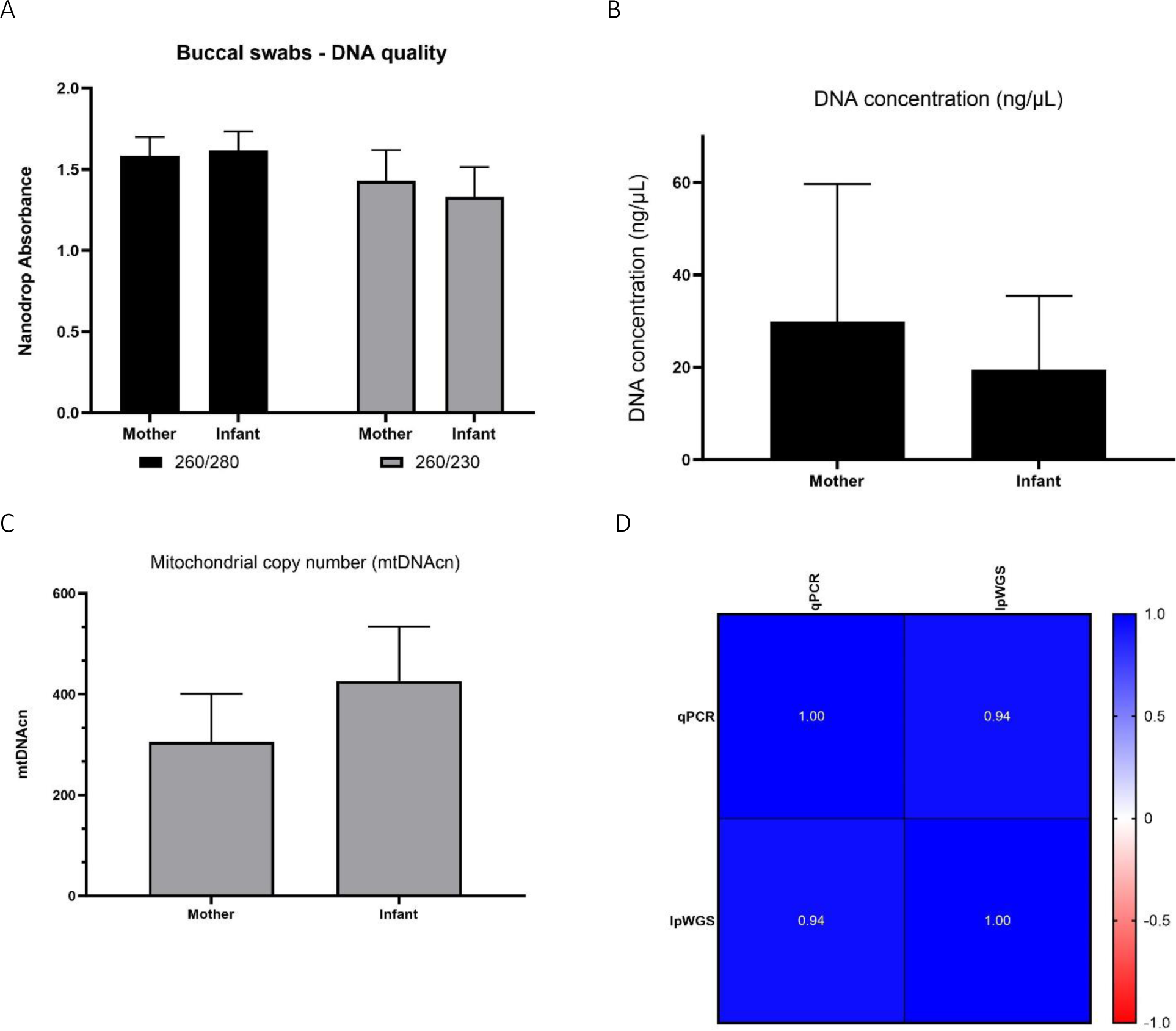
ACEs study: Nucleic acid collection results and mtDNAcn results from our qPCR and lpWGS assays. DNA quality (Panel A), DNA quantity (Panel B), average mtDNAcn in mothers (164) and infants (164) assessed using our qPCR assay (panel C), and concordance of 36 samples re-assessed for mtDNAcn using lpWGS (panel D), depicted as Pearson’s r values. RNA quality and quantity was also assessed and is shown in **Supplementary Figure s7**.

### 3.3 Mitochondrial haplogroup assessment

Mitochondrial haplogroups are defined by a set of variants in the mitochondrial genome that form specific branches of the mitochondrial phylogenetic tree (32, 33). These may be useful as markers of genetic ancestry, population migrations and environmental adaptation, and certain haplogroups have been associated with susceptibility or resistance to specific diseases (34, 35). We used the Phy-mer tool that we previously developed (33) to evaluate the mitochondrial haplogroups and determine haplogroup regions for 18 mothers for which we had lpWGS data (**Figure 6**). The majority of maternal haplogroups were from the “Americas” region (North and South America).

**Figure 6.**
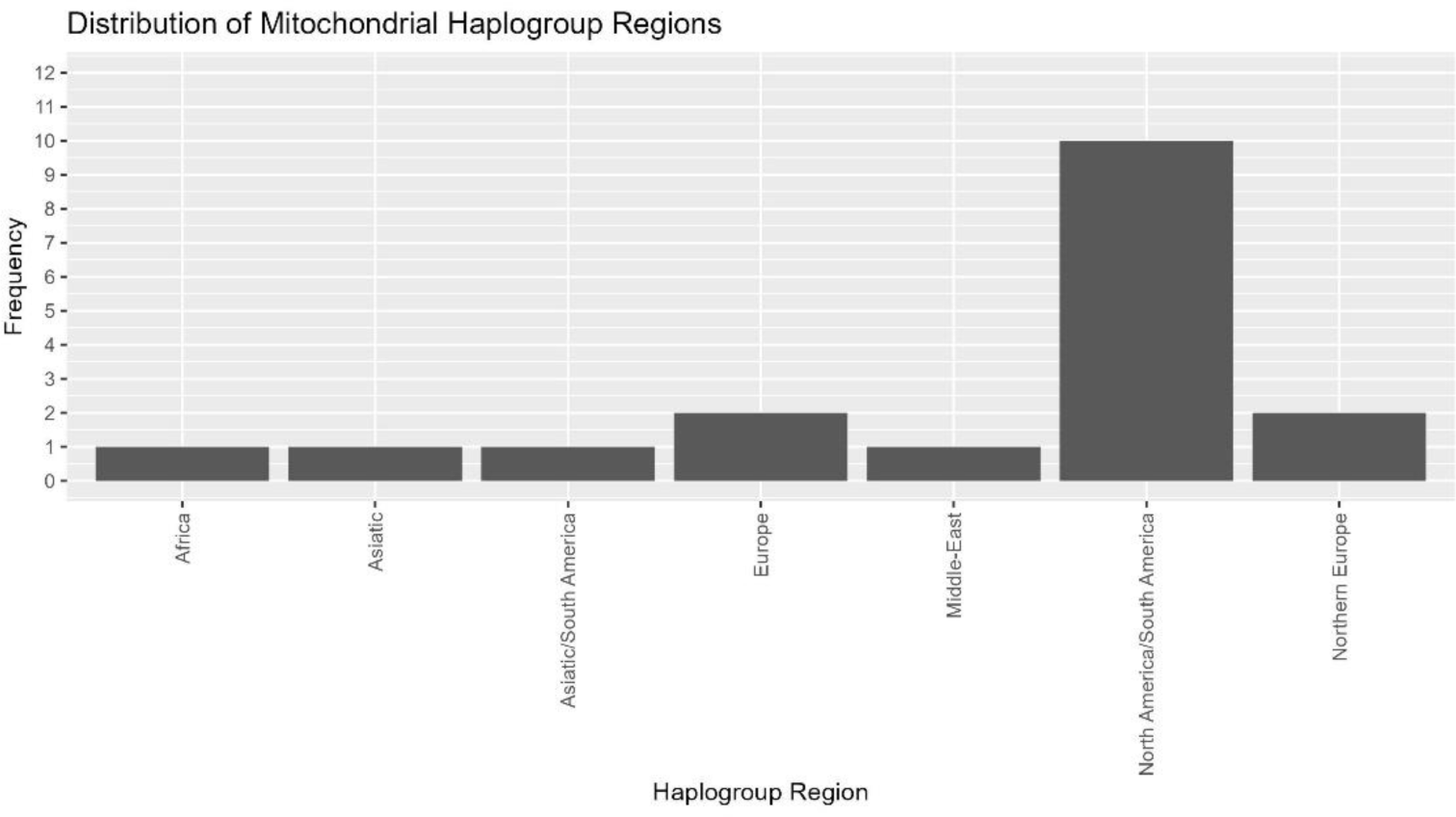
Mitochondrial haplogroups determination. Evaluation of mitochondrial haplogroups for 18 maternal samples that underwent lpWGS showed most falling into North America/South America, with small numbers from other haplogroup regions.

## 4 Discussion

We report the development and validation of mtDNA copy number assays that provide highly reliable and reproducible results from an oral biospecimen. Using the IsoHelix SK-2S buccal swab kit we were able to isolate DNA and RNA of sufficient quality and quantity from infants and adults to perform qPCR based mtDNAcn and lpWGS assays from a single sample. This makes these assays particularly useful for infants and young children for whom specimen collection and sample limitations may effectively preclude existing methods. Our own qPCR assay required as little as 2ng of DNA per qPCR replicate and our lpWGS required 5ng for library preparation. We demonstrated high concordance of results from our qPCR mtDNAcn assay, a commercial qPCR mtDNAcn assay, and lpWGS.

Different methods of measuring mtDNAcn have been evaluated previously (21). One study used data from two adult cohorts, the Atherosclerosis Risk in Communities (ARIC) study and the Multi-Ethnic Study of Atherosclerosis (MESA), and evaluated mtDNAcn calculated from qPCR, two microarray platforms, whole genome sequencing (WGS), and whole exome sequencing (WES) (21). The results showed that mtDNAcn calculated from WGS data had the strongest associations with known mtDNAcn correlates, such as age, sex, white blood cell count, and incident cardiovascular disease. mtDNAcn calculated from WGS also was more significantly associated with these traits compared to all other methods, including qPCR. While the current study only evaluated one DNA extraction method, the ARIC study also examined the impact of DNA extraction methods on mtDNAcn estimation and found that measuring mtDNAcn from cell lysate resulted in less variability compared to traditional methods using phenol-chloroform-isoamyl alcohol and silica-based column selection (21).

There is growing evidence that mitochondrial adaptation as part of the cell danger response (36, 37) may play a central role in various pathophysiological states, including cancer, cardiovascular disease, metabolic disorders and chronic stress (16, 19, 38–43). Mitochondrial DNA copy number is a critical determinant of mitochondrial function, influencing cellular energy production and metabolism (3). Developing mtDNAcn assays that can be used in infants and young children provides the opportunity to study whether atypical mtDNAcn may be an indicator of early pathophysiological changes that are currently challenging to identify using other measures. Use of buccal swabs for sample collection and a qPCR based method make our assay potentially cost effective and scalable, two key features for any clinical test. While this test has broad potential application, we are particularly interested in its utility for measuring mtDNAcn changes in infants and young children to better understand whether and how mtDNAcn may reflect developmental disruption due to early adversity. This requires study of larger populations which hopefully can be more readily accomplished using this assay.

## 7 Author Contributions

Simran Maggo and Dejerianne Ostrow led the development of the qPCR assays. Simran Maggo drafted the original manuscript including data analysis and production of tables and figures. Liam North and Aime Ozuna led the recruitment of study participants for the ACEs study. Yander Grajeda conducted all laboratory procedures to generate all data in this manuscript. Hesmedin Hakimjavadi assisted with lpWGS analysis. Jennifer Cotter, Alexander Judkins, Pat Levitt and Xiaowu Gai developed the ACEs study. Pat Levitt developed the Your Baby study. All authors contributed to editing and final proof-reading of this manuscript.

## 9 Acknowledgments

This research project was supported by the California Initiative to Advance Precision Medicine – ACES Program (OPR20139), The Saban Research Institute Developmental Neuroscience and Neurogenetics Program, the Simms/Mann Family Foundation and the CHLA Center for Personalized Medicine. We are grateful to Drs. Alma Gharib and Sahana Nagabhushan Kalburgi for guidance in establishing the full research protocol and to Suchi Patel for assisting with initial data generation for the NovaQUANT assay.

## Data Availability Statement

The original contributions presented in the study are included in the article/supplementary material, further inquiries can be directed to the corresponding author.

## 11 Supplementary Material

### 11.1 Inter and intra-day validation of the commercial NovaQUANT mtDNA qPCR assay

**Figure s1.**
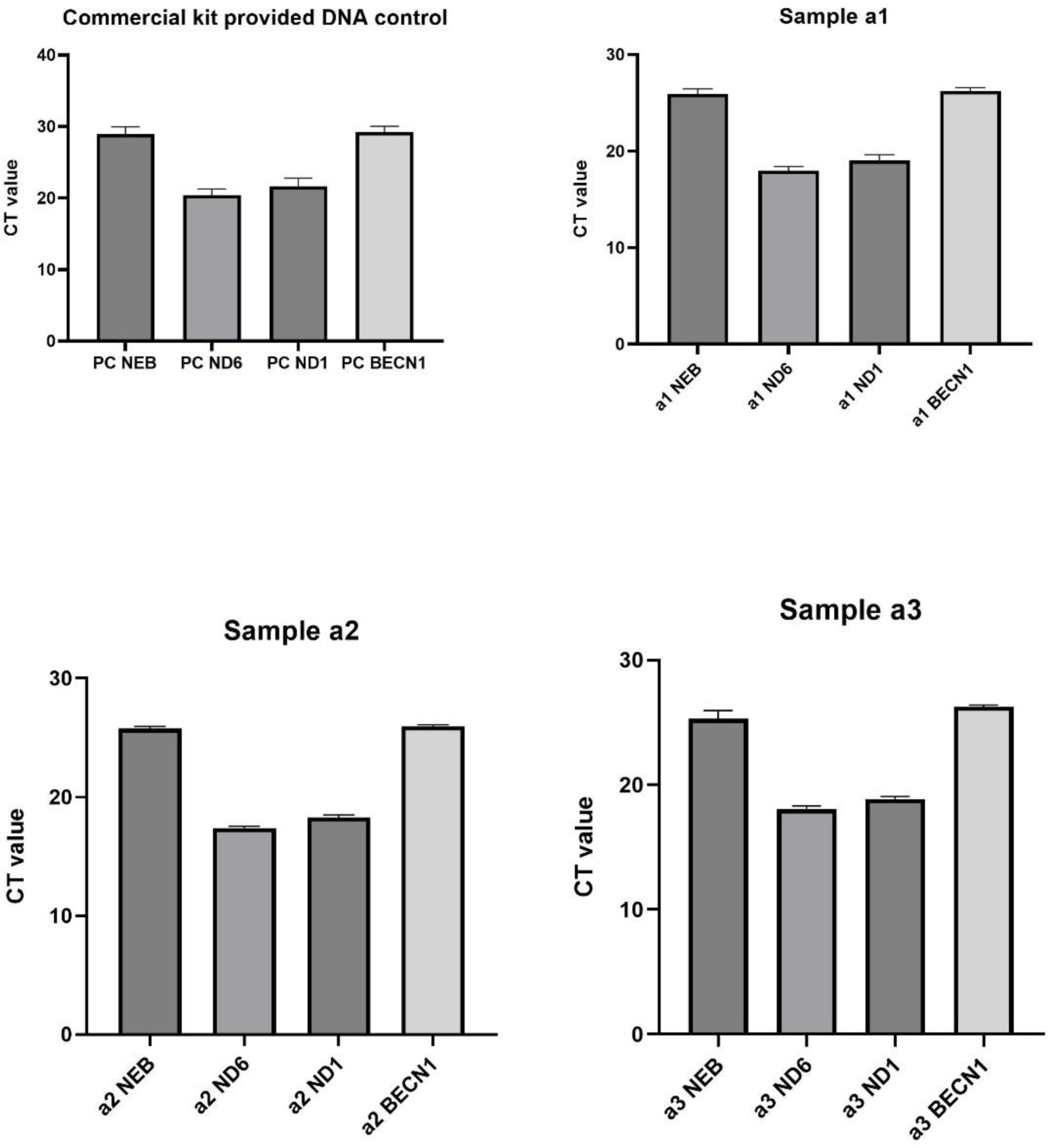
Mitochondrial DNA copy number assay validation. The qPCR assay utilizes two nuclear (NEB, BECN1) and two mitochodnrial genes (ND1, ND6). The graphs above show the average CT values ± SD from three separate validation runs. In each run the individual DNA sample was run in triplicate. Positive control (PC) is a DNA sample provided by the kit manufacturer.

**Figure s2.**
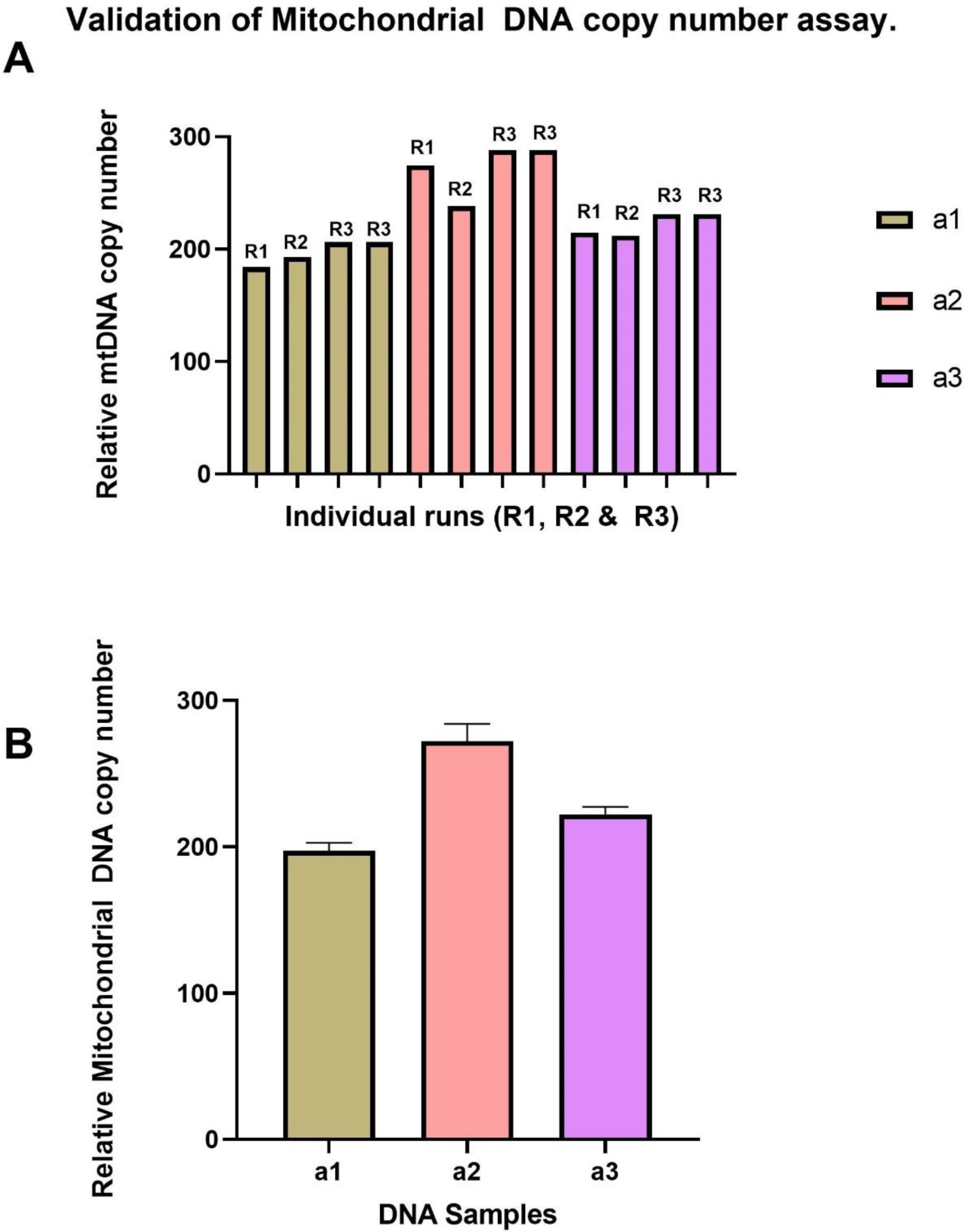
Panel A - DNA samples were assayed in 3 separate validation runs (R1, R2, R3). R1 and R2 are inter-day validation runs. R3 is the intra-day validation run. In the third validation run (R3,intra-day), each sample was run twice (in triplicate). Panel B - Average of all runs. The graphs above show the average mitochondrial DNA copy number ± SD.

**Figure s3.**
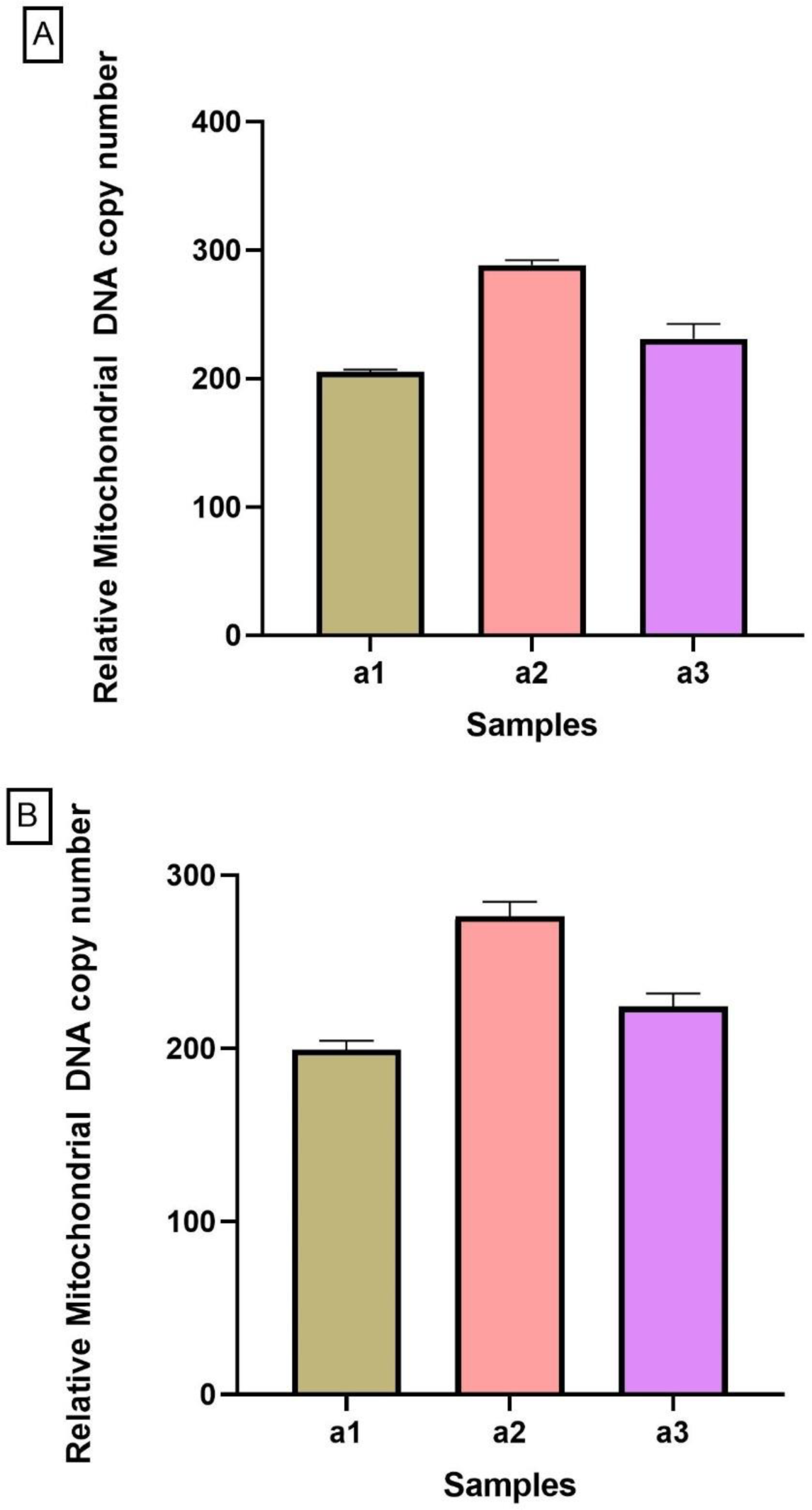
The graphs above show validation results from intra (A) and inter (B) runs. Inter-day results show relative mitochondrial DNA copy number ± SD from samples run consecutively on two separate days on different plates in triplicate. Intra-day results show relative mitochondrial DNA copy number ± SD from samples run TWICE on the same plate in triplicate.

### 11.2 Inter- and intra-day validation of our qPCR mtDNAcn assay

**Figure s4.**
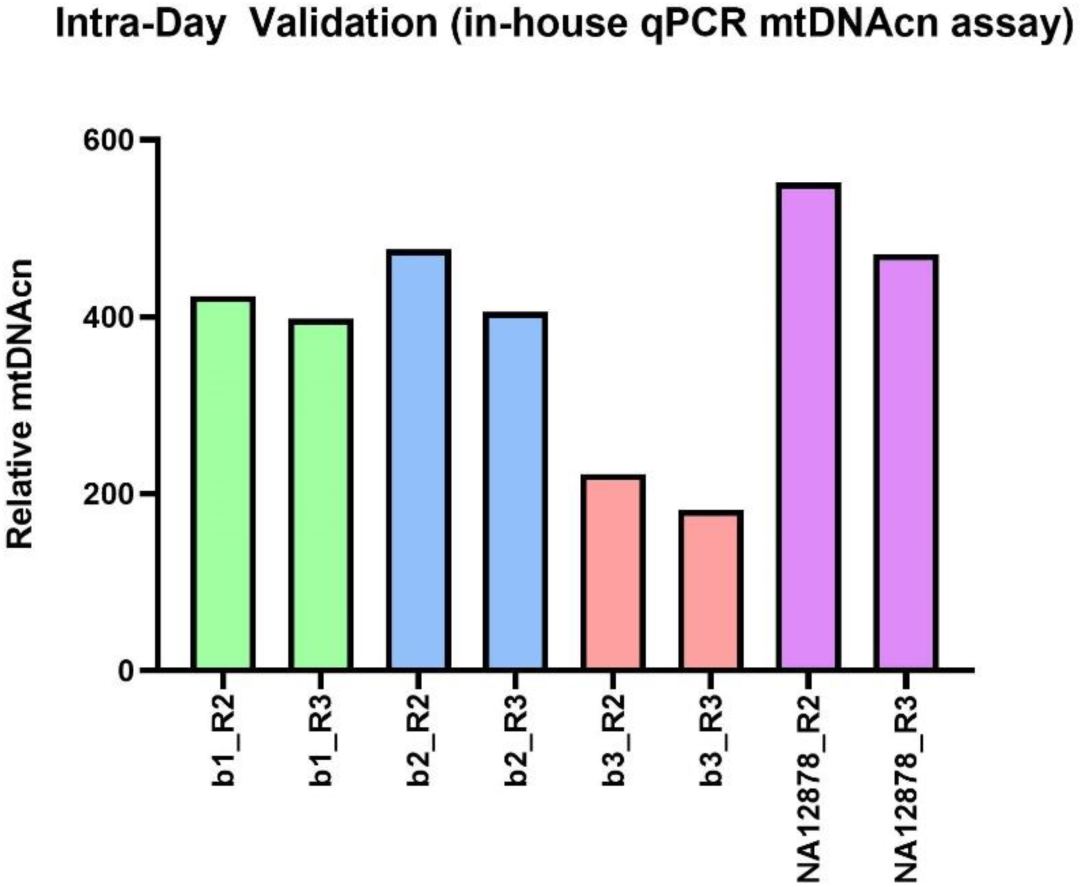
shows results of INTRA-day validation of in-house qPCR mtDNAcn assay. Run2 (R2) was carried out in the morning, and RUN3 (R3) was carried out in the afternoon of the same day. The different colored bars represent samples b1, b2, b3 and internal standard DNA sample - NA12878 respectively.

**Figure s5.**
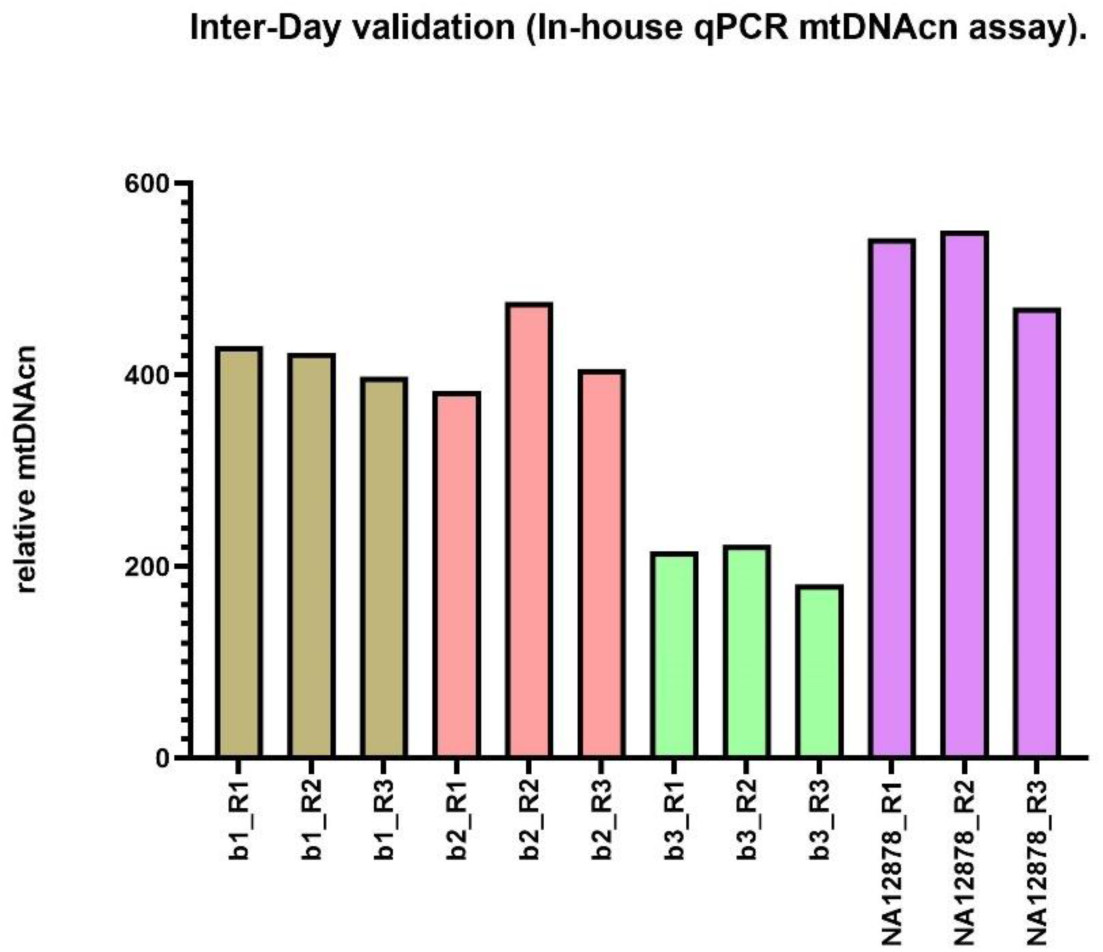
shows INTER-DAY validation of in-house qPCRmtDNA assay. Run1 (R1) and Run2 (R2) were carried out on consecutive days. Run 2 and Run3 (R3) were carried out on the same day. The different colored bars represent samples b1, b2, b3 and internal standard DNA sample - NA12878 respectively.

**Figure s6.**
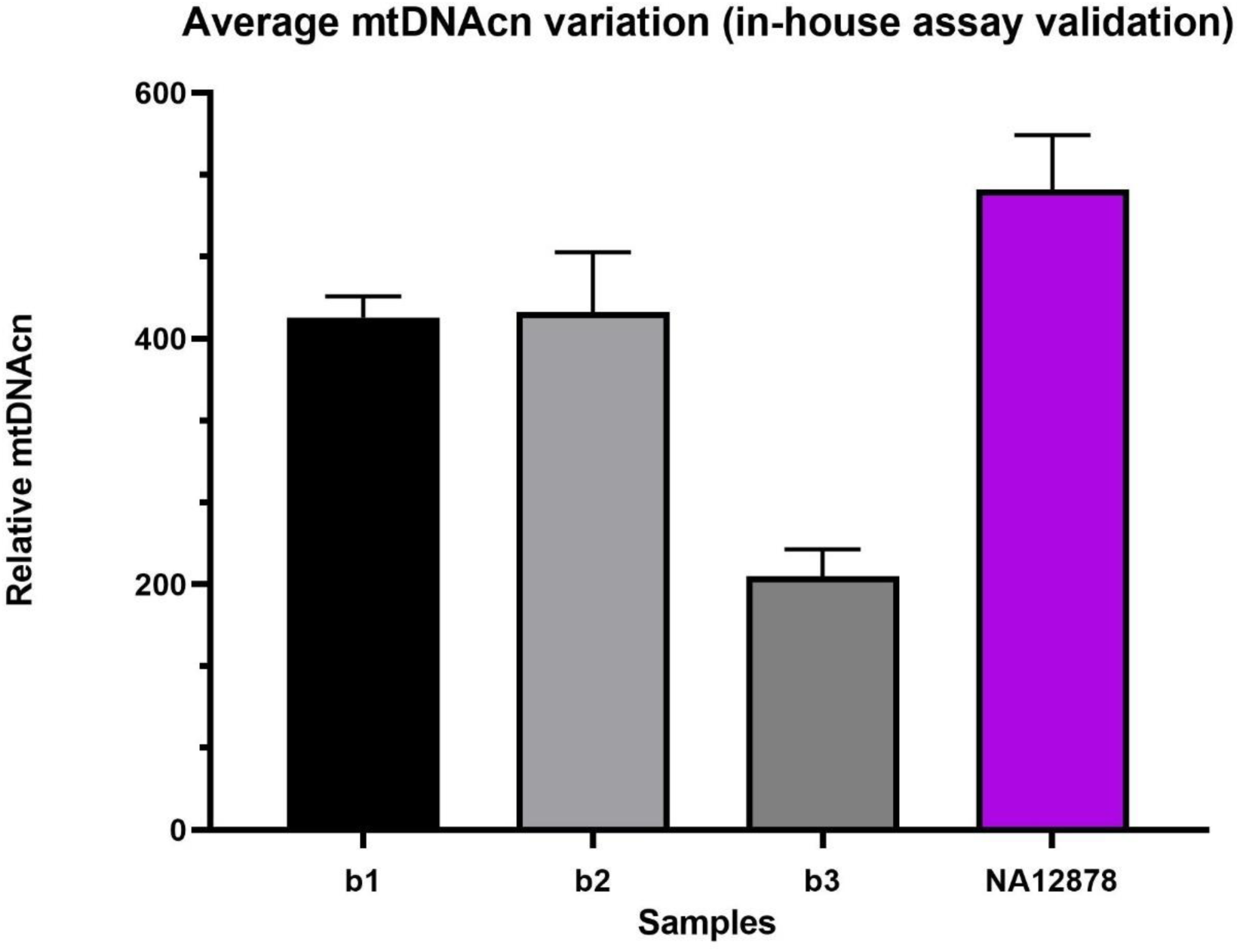
shows the average mtDNAcn and standard deviations from three separate runs.

### 11.3 RNA quality and quantity of extracted RNA from ACEs study

**Figure s7.**
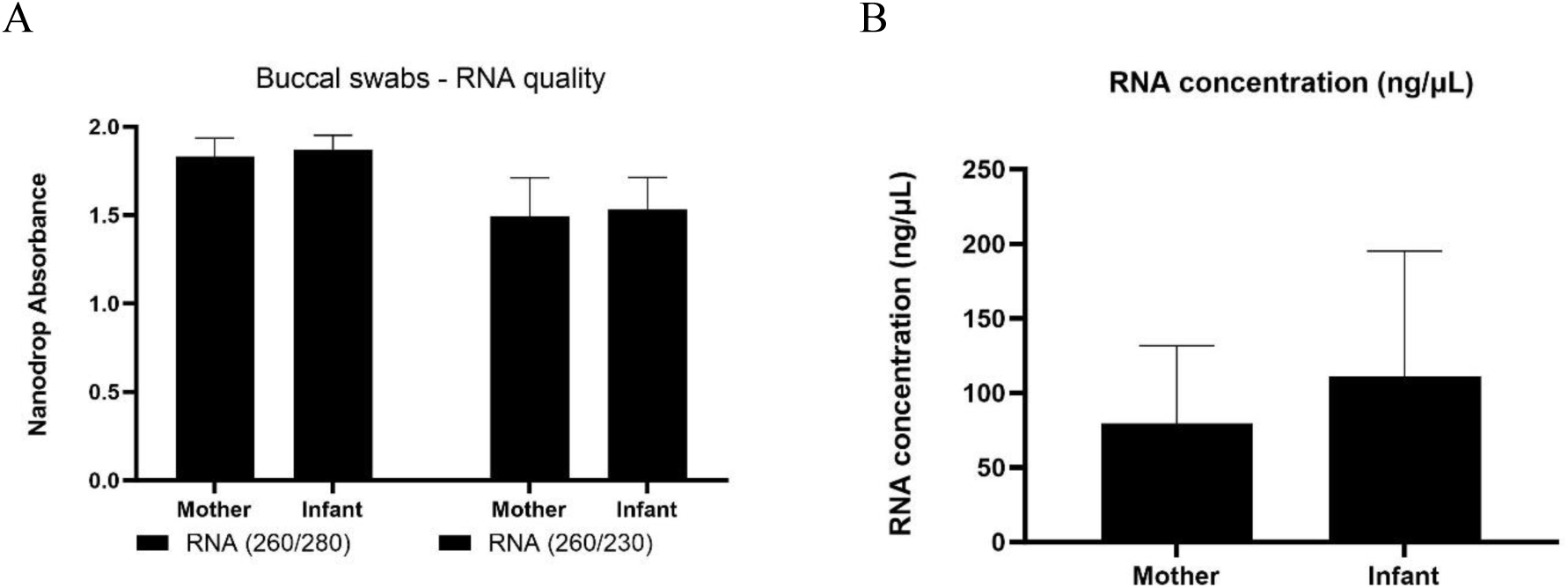
RNA quality (panels A) and quantity (Panel B) of 164 mother and 164 infant samples from the ACEs cohort.

